# Comparative evaluation of Oral lesions: “Tale - the Covid 19 Tells”

**DOI:** 10.1101/2022.02.03.22269712

**Authors:** Rashmi Bhavasar, Namratha A. Ajith, Rahul Bhavasar, Arti Shah, Vivek Vaswani

**Affiliations:** Department of Oral Pathology, KM Shah Dental College and Hospital, Sumandeep Vidyapeeth Deemed to be University, Piparia, Waghodia, Vadodara, Gujarat; Intern, KM Shah Dental College and Hospital, Sumandeep Vidyapeeth Deemed to be University, Piparia, Waghodia, Vadodara, Gujarat; Department of Pharmacology, Dr. Ulhas Patil Medical College and Hospital, Jalgaon, Maharashtra; Department of Pulmonary Medicine, Dhiraj Hospital, SBKS&MIRC, Sumandeep Vidyapeeth Deemed to be University, Piparia, Waghodia, Vadodara, Gujarat; Department of Medicine, Dhiraj Hospital, SBKS&MIRC, Sumandeep Vidyapeeth Deemed to be University, Piparia, Waghodia, Vadodara, Gujarat

**Keywords:** Oral Lesions, COVID 19, Sublingual varicosity, Candidiasis, Tongue

## Abstract

**Introduction & Objectives:** The COVID-19 pandemic has been raging across the globe since early January 2020. India has reported over 27 million cases and more than 3, 00,000 deaths. This study was planned to analyze the differences in demographic, clinical features and oral manifestations of COVID 19 patients hospitalized during COVID-19 pandemic.

**Methods:** This observational pilot study had total 36 participants, 12 each of mild, moderate and severe RT-PCR positive COVID cases hospitalized during COVID 19 pandemic. All demographic, clinical features, treatment details and oral manifestations were noted from first day of admission to hospital till treatment completion with follow up of minimum 7 days.

**Results:** Mean age of the patients was 39.44 ±9.13 years with M: F ratio of 5:4. Most common clinical presentation was fever, shortness of breath and treatment involved was symptomatic with supplemental oxygen & mechanical ventilation. Most common oral site involved was tongue & oral lesions observed were herpes labialis, mucositis, burning sensation, dryness of oral cavity, angular chelitis, aphthous ulcers, geographic tongue, fissuring of tongue, candidiasis, coated tongue, sublingual varicosity, & scalloped tongue.

**Interpretation and Conclusion:** All demographic, clinical and oral manifestations were significantly different in mild, moderate and severe cases of covid hospitalized patients. Though clinical symptoms were improved, oral lesions were worsened. Oral Lesions seen in covid patients were associated with multiple drug therapy for illness along with poor oral hygiene, but further etiology for lesions needs to be evaluated. Sublingual varicosity was observed in our hospitalised covid patients, but large sample observation is required for confirmation of findings and may be an early oral feature for covid detection. Prevention is always better than cure, so all patients positive for Covid should have a full mouth examination. Oral health should be priority during overall management of COVID patients and dentists should be a part of Covid management team.

## Introduction

As we all are aware, the COVID-19 pandemic has been raging across the globe since early January 2020. The appearance in December 2019, of a new corona virus has caused an unprecedented pandemic in the modern era. The novel corona virus disease (Covid 19) outbreak is still a pandemic affecting almost 240 million people worldwide with more than 4.8 million deaths. Surely, the disease produced by the novel coronavirus and its consequences have posed a challenge for health authorities worldwide. The ways of contagion through direct contact and through saliva in the form of small drops and the production of aerosols, & fomites have facilitated the rapid spread worldwide mainly infecting respiratory, gastrointestinal, & central nervous systems. Various geographical regions have been experiencing multiple waves of upsurge of cases which are not matched temporally as well as in severity.^1^ the first case of COVID -19 was reported in India on January 30, 2020 ^2^ since then, India has reported over 27 million cases and more than 3, 00,000 deaths^3^

Need of study: COVID-19 disease has distinct symptomatology outcome across the geographical borders. This leads to Restrictive actions for preventing the spread of the virus and limiting contagion (social isolation and self-quarantine). But this prevention poses immediate challenges for dentist. Comprehensive oral evaluation of covid lesions can lead to detailed insight of oral health, limiting its further spread. With this need of an hour, Study was planned by assuming Null Hypothesis that, “There is no difference or variation in oral health status and clinicopathological & oral symptoms during standard care of treatment in hospitalized covid positive patients with varying severity”

Aim of study was to evaluate and compare oral & clinical health in varying severity of covid patients hospitalized for standard care of treatment, comprising objectives as, first to evaluate oral health in varying severity of covid patients hospitalized for standard care of treatment., To evaluate clinicopathological parameters in varying severity of covid patients hospitalized for standard care of treatment, And To compare oral health and clinicopathological parameters in varying severity of covid patients hospitalized for standard care of treatment.

## Material and Methods

Place of study was - the Covid Unit of Dhiraj Hospital, India. Participants selected for the study were those hospitalized for treatment. Working Protocol & all guidelines using PPE kit was followed strictly while working in Covid unit under the guidance of supervising faculty of covid unit.

Participants in study were recruited using following Patient selection criteria for the study: Inclusion criteria for study were: Participants with age from 18 to 60years, RT-PCR Diagnosed covid positive cases hospitalised for standard care of treatment.

Whereas exclusion criteria were: Participants with any other major systemic illness at the time of admission to hospital, Participant who refuses to give a written consent, & Participants with history of tobacco abuse in any form

- This prospective observational pilot study had total of 36 participants, 12 in each mild, moderate & severe covid study group. Patient sample size was calculated using formula for sample collection for pilot study with 95% Confidence Interval and 80% power.

Ethical clearance for conducting study was obtained from the Institutional Ethics Committee (SVIEC). All Participants were informed about study through Participant’s information sheet. Once Participants were informed regarding study, they were recruited in the study only after taking their consent for study. Following consent, covid RT PCT positive patients were examined for oral lesions during their hospitalization for standard care for covid. Intra-oral examination included oral health status assessment using disposable mouth mirror and probe. Hard tissue as well as soft tissue observation was performed. If any lesion observed in the oral cavity, it was entered in proformas and photographs for the same were taken following their photographic consent. Covid sterilisation & disinfection protocol was strictly followed during patient interaction and data collection for study, as per Covid -19 Guides, & Confidentiality of study subjects was maintained. Following observations of the study, Descriptive statistics was performed, and data was subjected to statistical analysis by using (IBM) SPSS Version 20.0 Software.

## Observation & Results

1. **Herpetiform eruption comprising vesicles and erosions were seen in mild covid positive female on upper lip, 2 & 3 day of visit showed encrustations of the the lip lesion, which was completely healed by 7**^**th**^ **day.**
2. **Cracking, ulceration, white lesions and Depapillation of tongue mucosa along with lip dryness, ulceration, swelling of lips was present in moderate severity of covid positive patient**.
3. **Depapillation, sublingual varicosities of tongue in moderate whereas White lesions resembling curd like pseudomembranous candidiasis were seen on lower labial mucosa of mild covid positive patients**
4. **Central depapillation, geographic tongue, & peripheral whitish lesions on dorsal surface of tongue in moderate covid patients**.
5. **Hairy, fissured and yellowish, brown pigmented dorsal surface of tongue in moderate covid patients**
6. **White lesion involving marginal & attached gingiva and angular chelitis along with thick coated tongue was present in the same patient who was severe covid positive**.
7. **Depapillation of tongue and sublingual varicosity were observed in severe covid positive female patient**.
8. **Fissured tongue along with white membranous lesions and scalloping with teeth indentations on tongue were present in moderate covid positive patients**.
9. **Radiological investigations on chest x ray revealed Minimal costal pleural thickening and ground glass opacities**
10. **Consolidations and Multiple patches of ground glass density in sub pleural areas of lung**.
11. Microbial culture investigations from swab test of oral lesions revealed presence of round creamy white colonies on blood agar and microscopic evaluation showed presence of gram positive candida fungi.

Patient counselling for symptomatic Palliative care was done during examination for maintaining oral health and after discussion with physician antifungal treatment consisted of, Fluconazol, nystatin, and chlorhexidine 0.12% mouthwash, as oral lesions can be Secondary lesions resulting from deterioration of systemic health or due to treatments for COVID-19.

**Table 1A:**
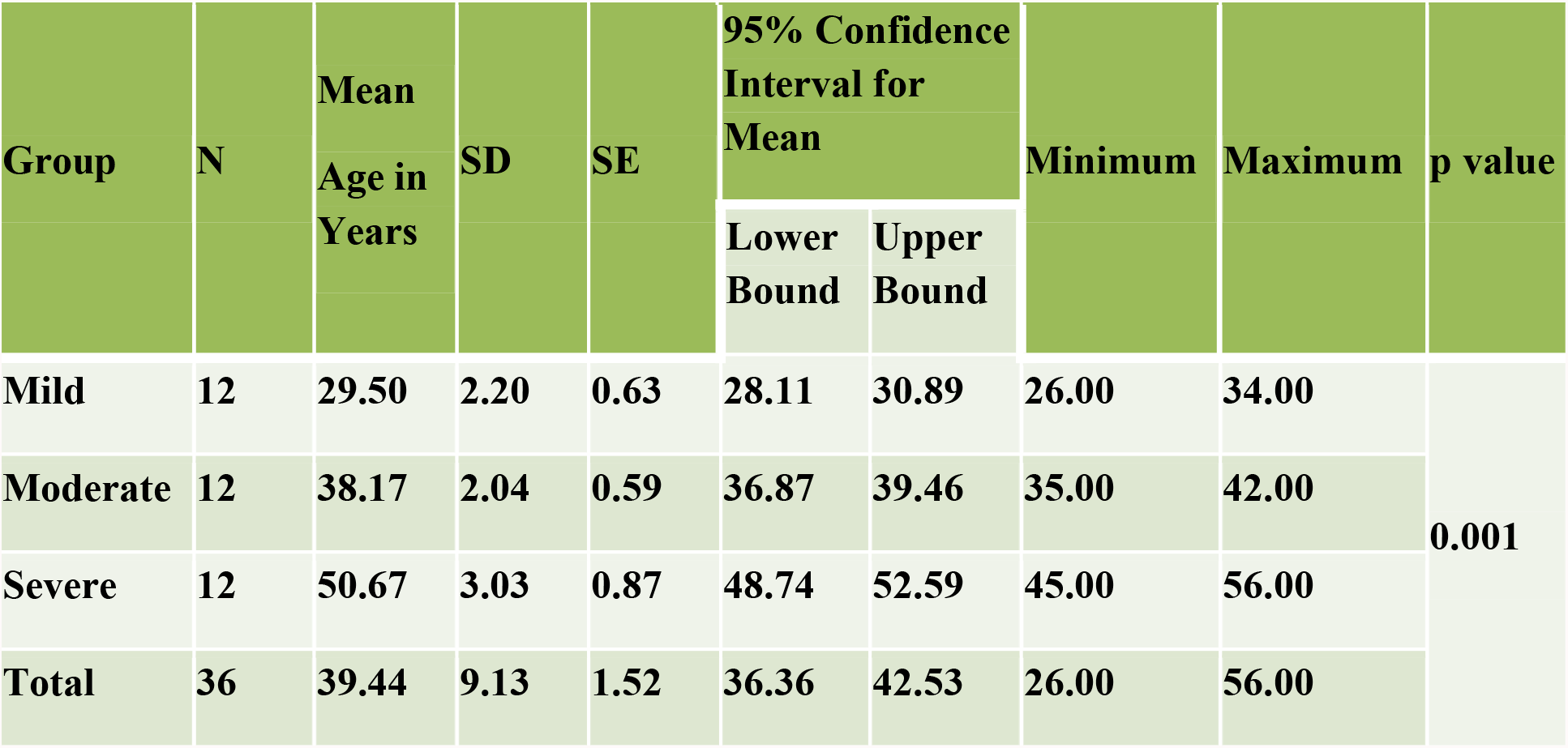
Age Distribution in Mild, Moderate and Severe Covid Patients:

**Table 1 & Graph 1.**
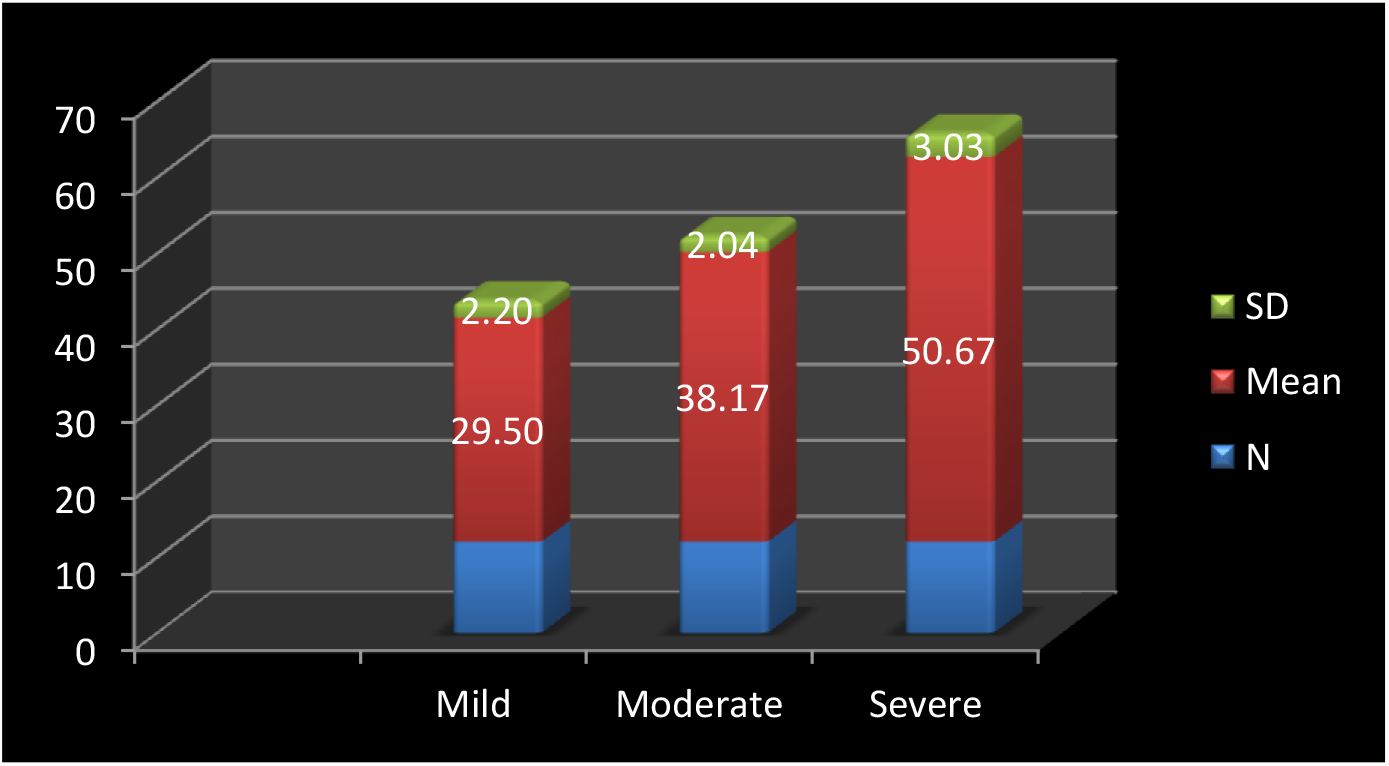
of Age Distribution in Covid Patients revealed, younger age involvement in mild group whereas older individuals were affected in severe group. Irrespective of severity categorization, mean age for participants was 39.44 years. The difference of age group in mild, moderate and severe groups of covid patients was statistically highly significant.(P< 0.001)

**Table 1B:** Age and Gender Distribution in Mild, Moderate and Severe Covid Patients:

| Severity | Gender | N | Mean | SD | p value |
| --- | --- | --- | --- | --- | --- |
| MILD | M | 7 | 29.14 | 1.77 | 0.571 |
|  | F | 5 | 30.00 | 2.83 |  |
| MODERATE | M | 9 | 38.22 | 1.99 | 0.903 |
|  | F | 3 | 38.00 | 2.65 |  |
| SEVERE | M | 4 | 48.25 | 2.22 | 0.041 |
|  | F | 8 | 51.88 | 2.70 |  |

M:F Ratio in our study was 5:4 showing more number of male participants in mild and moderate cases whereas in severe group females were more and was found to be statistically significant.(P=0.04)

**Table 2.**
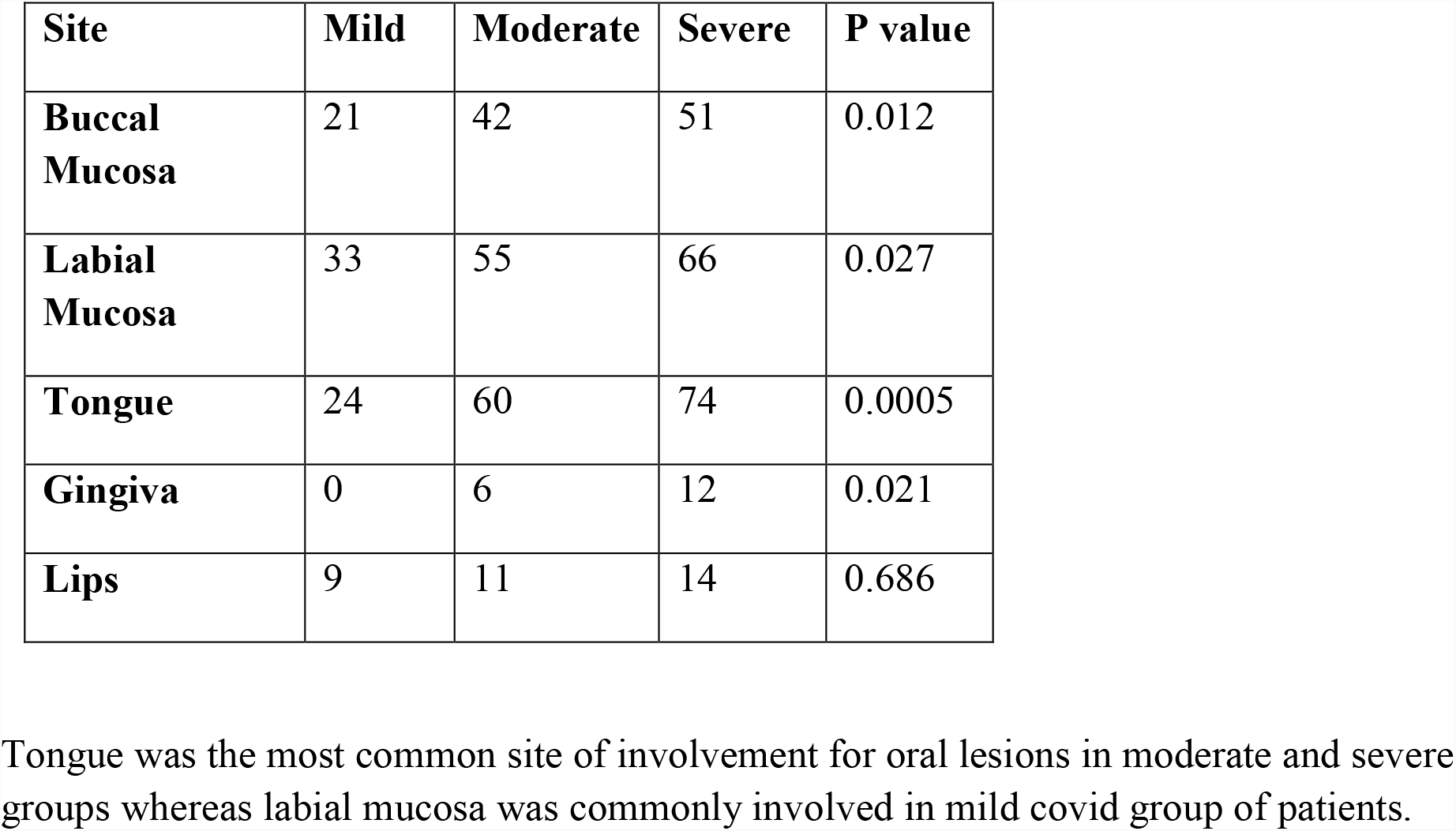
A: Site involvement for oral lesions in Mild, Moderate and Severe Covid Patients:

**Table 3:** Radiological presentation in Mild, Moderate & Severe Covid Patients:

| PARAMETERS | MILD | MODERATE | SEVERE |
| --- | --- | --- | --- |
| Radiological Presentation | Minimal costal pleural thickening | Consolidations with irregular septa<br>Scattered ground glass opacities | Multiple patches of ground glass density throughout lung.<br>Lymph adenopathy in precordial, Paratracheal regions. |
| Hospitalization Stay | 1 – 3 days | 5 –7 days | ICU- 7 – 10 days |

Hospitalization Stay was 1 to 3 days for mild whereas 7-10 days for severe cases.
Radiological Presentation on mild cases was Minimal costal pleural thickening whereas Consolidations with irregular septa & ground glass opacities were commonly seen in moderate and severe cases.

**Table 4:** Distribution of General Common symptoms in Mild, Moderate & Severe Covid Patients:

| Common complaints noted during hospitalization | Mild | Moderate | Severe |
| --- | --- | --- | --- |
| Anosmia | 8 | 12 | 12 |
| Fever | 12 | 12 | 12 |
| Sore Throat | 12 | 12 | 12 |
| Cough | 12 | 10 | 12 |
| Fatigue | 12 | 12 | 12 |
| Shortness of breath | 4 | 8 | 12 |
| Loss of appetite | 10 | 12 | 12 |
| Nausea, Vomiting | 8 | 10 | 12 |
| Headache | 10 | 12 | 12 |
| Myalgia | 10 | 12 | 12 |
| Nasal Obstruction | 11 | 12 | 12 |
| Diarrhea | 5 | 9 | 10 |
| Dysphagia | 7 | 10 | 12 |
| Face pain/heaviness | 10 | 11 | 12 |
| Abdominal Pain | 03 | 06 | 05 |
| Chest Pain | 0 | 0 | 4 |
| Haemoptysis | 0 | 0 | 3 |
| Depression | 9 | 12 | 12 |
| Numbness/Paresthesia | 0 | 6 | 10 |
| Runny nose | 9 | 11 | 10 |
| Sneezing | 10 | 11 | 12 |
| Conjunctivitis | 0 | 4 | 7 |
| Loss / alteration of taste | 10 | 12 | 12 |

Most Common General Symptoms noted during study were fever, sore throat, fatigue, followed by nasal obstruction, headache, loss or alteration of taste, & cough followed by other common symptoms. (Table 4 & Graph 2)

**Graph 2:**
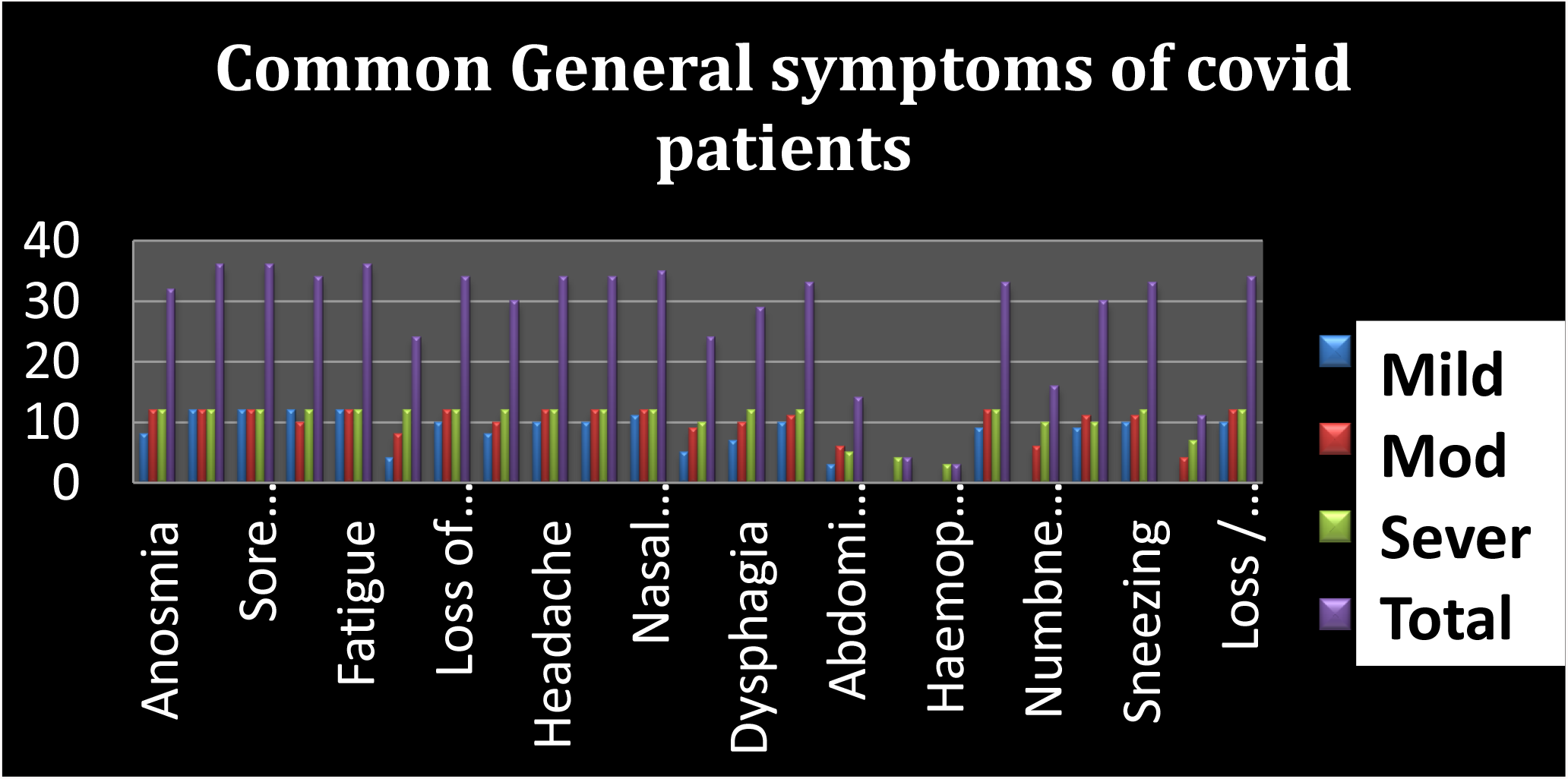
Common General Symptoms of Mild, Moderate & Severe Covid Patients:

**Table 5:** Distribution of oral lesions in Mild, Moderate & Severe Covid Patients:

| Common lesions noted during hospitalization | Mild | Moderate | Severe | P Value |
| --- | --- | --- | --- | --- |
| Herpes Labialis | 9 | 10 | 08 | 0.641 |
| Angular Chelitis | 06 | 08 | 11 | 0.083 |
| Aphthous ulcers | 05 | 10 | 09 | 0.212 |
| Lichen Planus | 00 | 01 | 02 | 0.754 |
| White plaque | 00 | 02 | 04 | 0.288 |
| Vesicles | 03 | 02 | 05 | 0.379 |
| Oedema of lips | 09 | 10 | 11 | 0.548 |
| Mucositis | 08 | 11 | 12 | 0.165 |
| Candidiasis | 02 | 08 | 12 | **0.0007** |
| Burning sensation | 06 | 10 | 12 | 0.172 |
| Geographic tongue | 01 | 05 | 04 | 0.165 |
| Fissured tongue | 02 | 07 | 06 | 0.091 |
| Multiple Yellowish Ulcers | 00 | 02 | 04 | 0.288 |
| Sublingual varicosity | 02 | 08 | 10 | **0.002** |
| Coated tongue | 06 | 04 | 10 | **0.042** |
| Dryness of mouth | 10 | 12 | 12 | NA |
| Tongue depapillation | 04 | 08 | 02 | **0.037** |
| Nodules of tongue | 00 | 01 | 00 | NA |
| Scalloped tongue | 01 | 05 | 10 | **0.001** |
| Ulceronecrotic Gingivitis | 00 | 00 | 02 | NA |
| Bleeding gingiva | 00 | 06 | 10 | **0.001** |
| Lesions due to medical devices | 00 | 01 | 03 | 0.940 |

**Graph 3:**
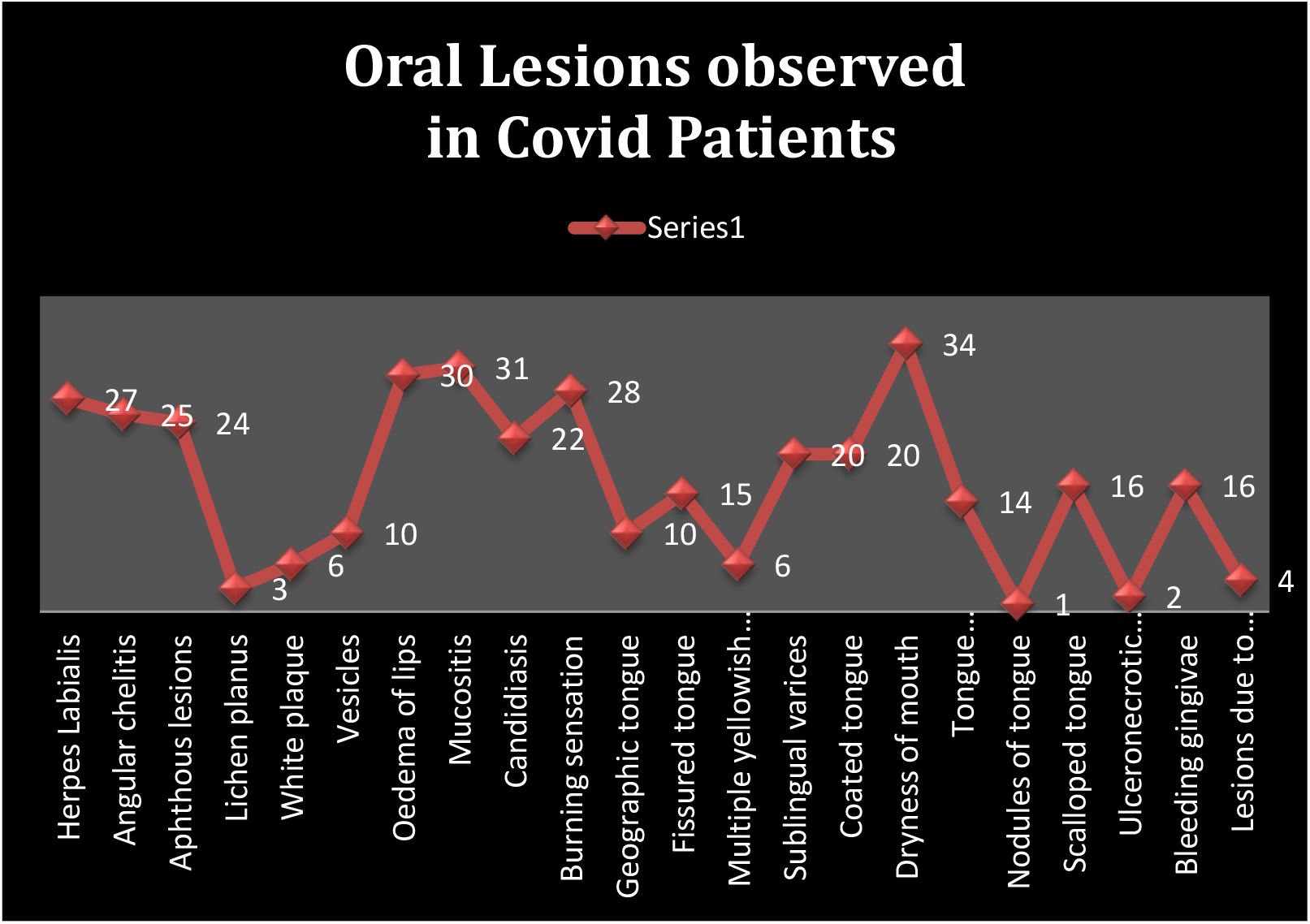
Distribution of Common oral manifestations seen in all mild, moderate and severe covid participants. Commonest feature observed was dryness of oral cavity followed by mucositis, edema of lips, burning sensation, herpes lesions followed by angular chelitis, candidiasis and coated tongue.

Oral health status in moderate & severe patients was poor whereas in mild cases, it was good.

## Discussion

Age distribution in mild, moderate and severe cases was varied but overall Mean Age of 39.44 ± 9.13 years in our study is younger than that reported by Tomo et al^4^ (50.5yrs) & M:F Ratio was 1:1, which was not in agreement with our study(M:F=5:4) as we found male participation more in mild & moderate cases but females were affected more in severe covid cases and was comparable to the studies reported so far but there are no studies reporting age as per severity of Covid cases. The most common general symptoms noted in our study were fever, sore throat, fatigue, followed by nasal obstruction, headache, loss or alteration of taste, & cough and was similar to those reported by Amorim Dos Santos J et al^5^ & by Chain et al^6^. The propable cause of common genral features is the the combined effects of COVID-19-related capillary damage, pre-existing microvascular changes, leads to vicious cycle, as infection- and hypoxia-related inflammation cause capillary function to deteriorate, which in turn accelerates hypoxia-related inflammation and tissue damage leading to its general symptoms in covid patients^7^

Among the distribution of site for oral lesions in our study, tongue both dorsal and ventral surfaces were followed by labial mucosa, buccal mucosa, gingiva and lips was commonly involved in mild covid group of patients which can be due to distribution of angiotensin converting enzyme 2 (ACE2) receptors & recently published tropism of SARS-COV-2 to the tongue and salivary gland epithelium suggest that the oral mucous membrane may be targeted by the virus as described by Xu, H., Zhong et al^8^.

Oral lesions seen in our study were in congruent with numerous oral lesions reported in patients with COVID-19 including a wide and varied range^5^

The most common specific oral symptom in all covid patients, found in our study in patients with COVID-19 was dryness of oral cavity, followed by mucositis, oedema, cracking of lips, loss of taste, burning sensation.

Dry mouth or xerostomia is a sign of dehydration which could occur secondary to underlying illness such as COVID-19 but viral induced infection and inflammation of salivary glands is a known cause of xerostomia as found by Saniasiaya, J^9^ who reported possible reason for xerostomia occurrence could be the neuroinvasive and neurotropism potential of SARS-Cov-2.

High grade fever of covid patients itself can lead to dehydration state of body, & salivary gland inflammation leading to gustatory impairment and thick ropy, viscous saliva is prone for growth of bacteria, fungi which in turn leads to halitosis, another common feature seen in our covid patients. Xerostomia can also be due to ventilation and oxygenated air for artificial oxygen requirement of covid patients if given with inadequate humidification itself becomes another reason for dryness, also patients were prohibited from water intake when on ventilation as requirement of covid treatment.

Our finding was in accordance to Study by Biadsee et al^10^ found 72 of their COVID-19 patients complained of xerostomia which reported a strong association with gustatory dysfunction and burning mouth. Alteration in the quantity and composition of saliva can lead to gustatory dysfunction. Loss of taste was also reported by Natto et al^11^ & Amorim Dos Santos J et al^5^ Anemia related to SARS-CoV-2 mediated hemolysis also causes the oral manifestations. ACE2, CD147, and CD26 receptors present on the erythrocytes are potential targets for SARS-CoV-2 attachment, which can lead to hemolysis. Loss of taste is also attributed to COVID -19 drug treatment side effects.

Oral lesions observed in our study were herpes lip lesions, coated tongue, candidiasis, fissured tongue, geographic tongue, viscous saliva, sublingual varicosity seen as specific oral lesions in COVID-19 patients. There is a possibility that the emotional distress of the situation itself could trigger such lesions in mild cases but in moderate and severe cases these lesions are more related to the drugs or immunosuppression. Immunosuppressed state can trigger a reactivation of the Herpes virus. A concomitant bacterial super infection may also occur due to reduced saliva or the lesions may arise from an inflammatory reaction that induces vascular inflammation. Gingival sulcus & periodontal pocket are favourable as a reservoir for the virus.

Various oral manifestation seen by various authors were in congruence with our study observations such as inflammation of the papillae of Wharton’s Duct hyposalivation ^12^, herpetiform eruption comprising vesicles and erosions on the lips ^13^, aphthous-like ulcers & persistent white plaque^14^, multiple pinpoint yellowish ulcers, nodule on the lower lip^15^ Various factors leading to oral lesions of covid patients are reviewed systematically^16,17,18^ but few major are, Immune system suppression, deteriorating general health, coinfections, opportunistic candidiasis, adverse reactions from medications to COVID-19 treatments, neglected oral hygiene, & drug hypersensitivity and urticaria may be related to the COVID-19 induced cytokine storm.

Moderate and severe covid cases in our study had poor oral health as compared to mild cases. Lesions are more related to stress, the drugs or immunosuppression driven by the drugs rather than to COVID-19.

Important oral finding noted in our study was occurrence of Sublingual varices seen as very characteristic and prominent small. Dilated veins under the lateral borders of the tongue. We had observed sublingual varicosity in 55.55% (20/36 cases) especially in moderate and severe cases and 2 cases of mild covid patients. Presence of sublingual veins is common clinical finding. Their pathogenesis has been reports may be due to a change in the connective tissue or weakening of the venous wall, as a result of degeneration of elastic fibers related to the ageing process, Their prevalence increases with age, & they are present in up to 60% of elderly patients, in both sexes, and in different population group.

Hedstorm et al^19^ found an association between sublingual varices and hypertension. It is found in association with conditions that result in increases venous pressure such as portal hypertension or superior venacava syndrome. Phleboliths or thrombophlebitis may complicate this condition.

Covid -19 has been described as vascular disease as SARS-CoV-2 leads to multi-system dysfunction with SARS-CoV-2-mediated endothelial injury, which is an important effector of the virus.^20^ It has also been related to thrombotic alterations produced by the virus itself. ^21^ Sublingual veins, though normal, its inflammation and increased prominence during covid needs to be observed and verified. Examining the lateral borders and sublingual veins of the tongue is easily done, and could be a valuable oral feature for the dental profession to take active part in preventive healthcare during covid. Thus, oral lesions can be an inaugural sign of Covid-19 or a warning sign of peripheral thrombosis.

## Conclusion

Significant differences were found on intergroup comparison of oral lesions in mild, moderate & severe covid groups. Identification of Oral lesions can significantly impact overall covid prognosis due to multiple secondary infections & influence treatment outcome. Oral Lesions seen in covid patients were associated with multiple drug therapy for illness along with poor oral hygiene, but further etiology for lesions needs to be evaluated. Sublingual varicosity was observed in our hospitalised covid patients, and may be an early oral feature of covid detection, as it has vascular pathogenesis. Prevention is always better than cure, so all patients positive for Covid should have a full mouth examination. Oral lesions could be the first COVID-19 signs to arise and hence dental practitioners are first to identify or suspect SARS-CoV-2-positive patients if any. Dental Faculty should be a part of Covid pandemic Care.

Limitation of our study was less number of patients with short follow up, also photograph of lesions was taken with transparent plastic cover on mobile for protection due to nature of the disease. Further studies should be planned with well distributed larger sample size and longer follow up to find out exact reasons of oral lesions for better treatment outcome.

## Data Availability

data details will be provided on request for verification

